# Acceptability and Feasibility of Combination Treatment for Cervical Precancer Among Women Living with HIV in South Africa: Primary Outcomes from the ACT 2 Randomized Trial

**DOI:** 10.64898/2026.03.14.26348308

**Authors:** Nicholas S. Teodoro, Katie R. Mollan, Jessica R. Keys, Chuning Liu, Masangu Mulongo, Sibusisiwe Gumede, Tafadzwa Pasipamire, Mark Faesen, Melissa A. Mischell, Lisa Rahangdale, Carla J. Chibwesha

## Abstract

**Objective:** Determine acceptability and feasibility of loop electrosurgical excision procedure (LEEP) combined with adjuvant intravaginal 5-fluorouracil (5FU) for treatment of cervical intraepithelial neoplasia grade 2/3 (CIN2/3) in women living with HIV (WLWH).

**Design:** Double-blind, randomized, placebo-controlled Phase 2b feasibility trial.

**Setting:** Public-sector hospital in Johannesburg, South Africa.

**Population:** 180 WLWH aged 18+ years with CIN2/3 confirmed by LEEP and on antiretroviral therapy for ≥60 days.

**Methods:** Participants underwent LEEP and were randomly assigned (1:1) to receive 8 doses of adjuvant 5FU or placebo cream every other week and followed for 24 weeks.

**Main Outcome Measures:** The primary outcomes were acceptability and feasibility (adherence, retention, safety, tolerability).

**Results:** Between March 2023 and January 2025, we randomized 180 WLWH. Median age was 41 years (interquartile range [IQR]: 35–45), median CD4+ count was 636 cells/mm³ (IQR: 376–873), and 98.9% were virologically suppressed.

Acceptability (>94%) and adherence (>91%) were high and comparable between arms. Retention exceeded 92% in both arms, although Week 24 attendance was lower in the 5FU arm (92.2% vs. 98.9%, probability difference [PD] -6.7%, 95% confidence interval [CI] -14.4%, -0.5%). Safety events were mild, more common with 5FU, and primarily reported as Grade 1 or 2 cervical inflammation (49.2% vs. 26.7%, risk difference [RD] 22.5%, 95% CI 8.6%, 36.4%). One Grade 3 adverse event (an allergic reaction to 5FU) resulted in treatment discontinuation.

**Conclusions:** LEEP plus adjuvant intravaginal 5FU is acceptable and feasible among WLWH in South Africa, supporting progression to a Phase 3 trial.

**Clinical Trial Registration:** NCT05413811.

**Funding:** United States National Institutes of Health (R01CA250850).

## Introduction

Cervical cancer is among the most common cancers in women, with over 90% of cases and deaths occurring in low- and middle-income countries (LMICs).^1, 2^ African women experience the highest burden of cervical disease due to disparities in prevention and treatment.^2^ The HIV epidemic in Africa also has a significant impact on cervical disease, with high-risk human papillomavirus (hrHPV) infection, high-grade cervical intraepithelial neoplasia (CIN2/3), and cervical cancer occurring more commonly in women living with HIV (WLWH).^3, 4^

For WLWH, treatment options for CIN2/3 remain suboptimal. Current management of CIN2/3 is based on surgery alone, most commonly loop electrosurgical excision procedure (LEEP) or ablation (e.g., cryotherapy or thermocoagulation).^5^ Surgical therapy is highly successful in women without HIV, failing in only 5–10% of cases.^6, 7^ By contrast, WLWH experience high rates of persistent/recurrent disease (20–55%), increasing their risk of progression to cervical cancer.^8–14^ Managing treatment failure requires repeat surgical procedures and close clinical follow-up, both frequently unavailable in LMIC settings.

Topical 5FU is a biologically plausible candidate for adjuvant treatment of CIN2/3. Its primary mechanism—inhibition of thymidine synthesis and DNA replication—is well suited to HPV-driven dysplasia. Topical 5FU is approved for dermatologic precancer and widely used off-label for HPV-associated vulvar, vaginal, and anal precancer.^15–17^ Importantly, two U.S. randomized trials have provided proof-of-concept for intravaginal 5FU in cervical disease: one demonstrated a significantly lower risk of recurrent CIN2/3 when 5FU cream was used following surgical treatment in WLWH,^18^ and a second showed higher rates of disease regression with 5FU compared to observation in immunocompetent women with CIN2.^19^

Despite this evidence, intravaginal 5FU has never been evaluated as adjuvant therapy for CIN2/3 in an African or broader LMIC setting, where the unmet need is greatest. To address this gap, we conducted a Phase 2b feasibility trial (“ACT 2”) of LEEP combined with adjuvant intravaginal 5FU cream in South African WLWH. Our primary objective was to determine the acceptability and feasibility (adherence, retention, safety, tolerability) of combination treatment for CIN2/3 among WLWH. Our secondary objectives (reported separately) were to assess the efficacy of combination treatment, including (a) regression of cervical disease and (b) clearance of hrHPV.

## Methods

### Trial Design, Randomization, and Masking

ACT 2 was a double-blind, randomized, placebo-controlled Phase 2b trial designed to assess acceptability and feasibility, rather than to test efficacy. Detailed study procedures were published previously.^20^ Briefly, WLWH with CIN2/3 confirmed on LEEP were enrolled in the trial and randomly assigned (1:1) to self-administer 8 doses of intravaginal 5FU cream or placebo cream (once every 2 weeks). Participants were followed for 24 weeks to assess acceptability and feasibility (adherence, retention, safety, tolerability) of combination treatment.

The randomization schedule used a permuted block design with randomly varying block sizes of 2, 4, and 6, and was created centrally to ensure allocation concealment. Treatment assignment was carried out by an unblinded pharmacist.

### Population

Women were recruited from the cervical cancer prevention program within an HIV clinic at a large, public-sector teaching hospital in Johannesburg, South Africa. Enrollment began in March 2023. Inclusion criteria were confirmed HIV-1 infection, age ≥18 years, receipt of antiretroviral therapy (ART) for ≥90 days, and an indication for LEEP based on biopsy-confirmed CIN2/3. In April 2023, the eligibility criteria were amended to an indication for LEEP based on high-grade cytology or biopsy-confirmed CIN2/3 and ART for ≥60 days. Participants were also required to use dual contraception during the study. Key exclusions were pregnancy, breastfeeding, active sexually transmitted infection (STI; women were eligible to participate after treatment), surgically absent cervix, prior anogenital cancer, biopsy-confirmed cervical cancer, and any comorbidity precluding participation.

### Intervention

LEEP was performed in accordance with the standard of care. The 5FU (Efudix) cream was supplied by a South African pharmaceutical company. An inert emollient matching the appearance and consistency of the 5FU cream was used as the placebo. Participants self-administered 2g of 5% 5FU or placebo cream intravaginally once every 2 weeks for a total of 8 doses.

### Procedures

Women interested in participation provided written informed consent and underwent screening (Week 0), which included medical history, confirmation of HIV infection, pregnancy and STI testing, pelvic examination, colposcopy, and LEEP. Additional baseline testing included Xpert HPV testing (GeneXpert System, Cepheid, Sunnyvale, California), CD4+ cell count, and HIV-1 viral load. Women attended a results visit at Week 2; those who did not meet study eligibility criteria were considered screen failures. Women diagnosed with STIs received treatment prior to enrollment.

On enrollment at Week 4, pregnancy testing, pelvic examination, colposcopy, and HPV testing were performed. Eligible participants with CIN2/3 confirmed on LEEP histology were randomly allocated (1:1) to receive 8 doses of intravaginal 5FU or placebo cream. Women received instructions on how to use the 5FU/placebo cream (Supplementary Material) and administered the first dose under observation. Subsequent doses were self-administered at home once every 2 weeks.

Follow-up visits occurred at Weeks 6, 10, and 18 and included an assessment of adverse events (AEs). Adherence to the study intervention was also reinforced. At the exit visit (Week 24), participants underwent pregnancy testing, pelvic examination, colposcopy, cervical biopsy, and, if indicated, endocervical curettage (ECC). STI testing was repeated, and samples were collected for cytology, HPV testing, CD4+ cell count, and HIV-1 viral load.

### Laboratory Testing

Pathologists blinded to study arm assignment reviewed the histology and cytology specimens. HPV and STI testing for *N. gonorrhoeae*, *C. trachomatis*, and *T. vaginalis* was performed using the GeneXpert system.^21–25^ We also used the Syphilis BD Macro-Vue (Abbott Laboratories, Abbott Park, Illinois) to screen for *T. pallidum* antibodies.^26^

CD4+ cell counts were determined by flow cytometry (FACSCanto Clinical Flow Cytometry System, BD Biosciences, Franklin Lakes, New Jersey). Plasma HIV viral load was measured using the Abbott RealTime HIV-1 Assay (Abbott m2000 system, Abbott Laboratories, Abbott Park, Illinois).

### Outcome Measures

Our primary outcomes were acceptability and feasibility, the latter assessed through adherence, retention, safety, and tolerability. Our secondary outcomes (reported separately) included regression of cervical disease to CIN1 or normal histology at 24 weeks and genotype-specific clearance of hrHPV at 24 weeks. These outcomes were selected to inform the design and implementation of a future Phase 3 trial.

Acceptability was measured using a 7-item Likert-scale questionnaire and reported as a percentage score (0–100%). Participants recorded cream use and side effects in study diaries. Women returned all used and unused 5FU/placebo applicators at follow-up visits. Adherence was assessed using applicator counts, ultraviolet inspection (UVI), and self-report, with UVI designated as the primary adherence measure. Participants were classified as adherent if they applied ≥6 of 8 doses. Retention was defined as completion of study follow-up through Week 24.

Safety assessments were conducted at Weeks 4, 6, 10, 18, and 24, and included pelvic examination and colposcopy. Safety events included study drug–related Grade 1 genital lesions (blisters, pustules, or ulcers) and any study drug–related Grade ≥2 AEs. Dose-limiting toxicities (DLT) were defined as study drug-related Grade 1 genital lesions or Grade ≥2 AEs resulting in dose reduction or drug discontinuation. Tolerability was defined as being able and willing to apply ≥4 of 8 doses of study cream. If the study drug was discontinued, women remained in follow-up.

### Statistical Analysis

Descriptive statistics were used to summarize baseline characteristics and assess clinically meaningful imbalances between the randomization arms. Primary analyses were conducted using a modified intention-to-treat (mITT) population, consisting of all randomized participants who received at least one dose of study drug. Per-protocol and intention-to-treat analyses were also performed (data not shown).

Acceptability scores were summarized descriptively and compared between arms at Weeks 10 and 24 using Wilcoxon rank-sum tests. The proportion of participants with an acceptability score ≥80% was estimated within each arm, and differences between arms were reported as probability differences (PDs) with 95% confidence intervals (CIs).

Adherence, retention, safety, and tolerability outcomes were analyzed as binary endpoints. For each outcome, proportions were estimated within each treatment arm with corresponding 95% CIs and compared between arms using PDs or risk differences (RDs) with 95% CIs, as appropriate. Agreement between adherence measures was assessed using the intra-class correlation coefficient (ICC). Kaplan–Meier methods were used to estimate time-to-event outcomes for study dropout and safety events, with the time origin as randomization (Week 4). Analyses were performed using R version 4.4.2 and SAS Enterprise Guide version 8.6.

### Sample Size

Our target sample size of 180 participants enrolled (90 per arm) was selected to provide precision for our primary outcomes. Safety and tolerability events were anticipated to be uncommon,^19^ and thus sample size calculations focused on the acceptability, adherence, and retention endpoints, with secondary consideration of safety and tolerability. We assumed 90% retention over 24 weeks, incorporating 10% attrition into all calculations.

Assuming 80% of WLWH would achieve an acceptability score ≥80% at Weeks 10 and 24, enrolling 180 women provides ±8.7% margin of error (MOE) within each study arm. Likewise, assuming 80% of WLWH would achieve adequate adherence (at least 6 of 8 doses), 180 participants provide an anticipated ±8.7% MOE for each study arm. With 90% retention over 24 weeks, the anticipated 95% CI precision spans 85%–94% pooled over arms and 82%–95% within arm. For safety and tolerability, 180 WLWH enrolled also yields acceptable precision (within arm: ±6.5% MOE for a 10% safety event proportion and ±4.7% MOE for a 5% tolerability event proportion).

### Data Safety and Monitoring

The trial was monitored externally and the data reviewed by an independent Data Safety and Monitoring Committee (DSMC) convened by the Lineberger Comprehensive Cancer Center at the University of North Carolina at Chapel Hill. The DSMC assessed accrual, retention, and safety. The committee also considered new scientific developments relevant to participant safety or the ethics of the trial.

### Ethical Approval and Clinical Trial Registration

The trial protocol was approved by the University of Witwatersrand Human Research Ethics Committee (201122) and the University of North Carolina at Chapel Hill Institutional Review Board (20-3565). The trial was registered with the U.S. National Library of Medicine at ClinicalTrials.gov (NCT05413811) and on the South African National Clinical Trials Register.

### Patient and Public Involvement

Educational materials for the trial (Supplementary Material) were co-designed with WLWH who participated in our pre-trial formative study.^27^ The results of the trial will be shared through local dissemination meetings and scientific conferences.

## Results

Between March 2023 and January 2025, we enrolled and randomized 180 WLWH with CIN2/3, meeting our target sample size. In total, we screened 221 women, and of these, 35 women did not meet inclusion criteria and 6 declined participation. Eligible WLWH were randomly assigned 1:1 to receive adjuvant 5FU cream (n=90) or placebo cream (n=90). All randomized participants received at least one dose of the study drug and thus our modified intention-to-treat analysis was equivalent to the intention-to-treat analysis (Figure 1).

**Figure 1.**
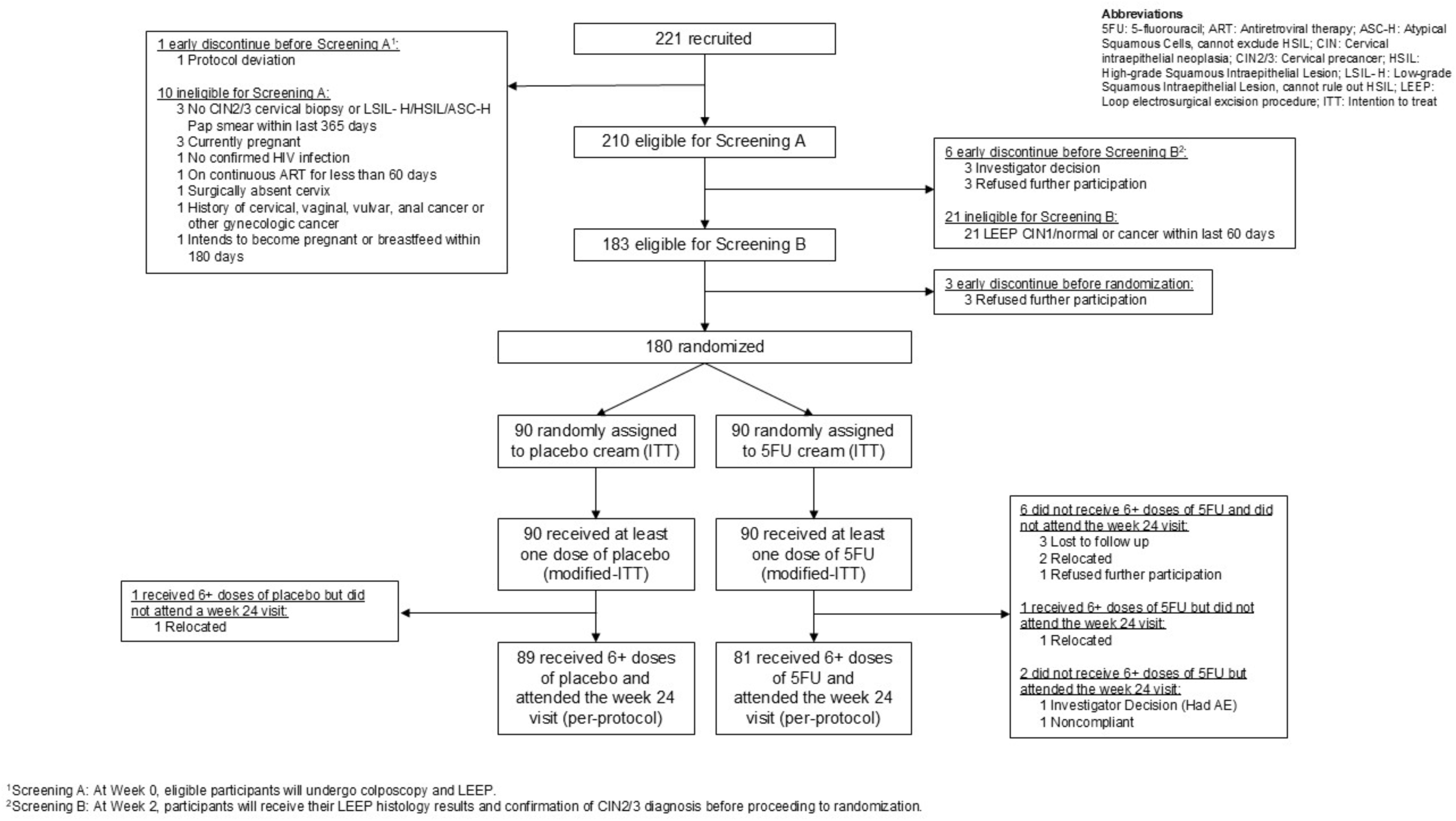
Participant Flow Diagram

Baseline demographic and clinical characteristics were similar between treatment arms (Table 1). Overall, median age was 41 years (interquartile range [IQR] 35–45), median CD4+ count was 636 cells/mm³ (IQR 376–873), and 98.9% (176/178) of women were virologically suppressed (HIV-1 RNA <200 copies/mL). Tobacco use was high (18.3%), as was the number of women diagnosed with STIs during screening (Trichomonas 10.6%, Chlamydia 6.7%, Syphilis 5.0%, Gonorrhea 3.3%).

**Table 1.** Baseline Characteristics of the Trial Population.

| Characteristic | Screened, Not Randomized (n=41) | Randomized |  |  |
| --- | --- | --- | --- | --- |
|  |  | Overall (n=180) | Placebo (n=90) | 5FU (n=90) |
| Demographics, median (Q1, Q3) or n (%) |  |  |  |  |
| Age, years | 42 (36, 48) | 41 (35, 45) | 42 (36, 46) | 39 (34, 44) |
| Time since HIV diagnosis, years | 9 (6, 14) | 9 (3, 14) | 8 (3, 13) | 10 (4, 14) |
| CD4+ cell count, cells/mm³ | 658 (438, 932) | 636 (376, 873) | 636 (398, 853) | 632 (342, 898) |
| HIV viral load |  |  |  |  |
| <200 copies/mL | 28 (97%) | 176 (99%) | 88 (99%) | 88 (99%) |
| ≥200 copies/mL | 1 (3%) | 2 (1%) | 1 (1%) | 1 (1%) |
| Sexual partners in the past year |  |  |  |  |
| 0 | 8 (24%) | 33 (18%) | 18 (20%) | 15 (17%) |
| 1 | 24 (71%) | 127 (71%) | 61 (68%) | 66 (73%) |
| 2 or more partners | 2 (6%) | 19 (11%) | 10 (11%) | 9 (10%) |
| Tobacco use in the past month |  |  |  |  |
| Yes | 10 (29%) | 33 (18%) | 18 (20%) | 15 (17%) |
| No | 24 (71%) | 147 (82%) | 72 (80%) | 75 (83%) |
| Cervical Disease Pre-study and at Baseline, n (%) |  |  |  |  |
| Pre-study Pap smear¹ |  |  |  |  |
| HSIL | 21 (70%) | 124 (80%) | 59 (76%) | 65 (84%) |
| ASC-H | 5 (17%) | 24 (15%) | 14 (18%) | 10 (13%) |
| LSIL or lower | 4 (13%) | 7 (5%) | 5 (6%) | 2 (3%) |
| Pre-study cervical biopsy¹ |  |  |  |  |
| CIN2 | 13 (54%) | 58 (41%) | 32 (47%) | 26 (36%) |
| CIN3 | 11 (46%) | 82 (59%) | 36 (53%) | 46 (64%) |
| **LEEP histology** |  |  |  |  |
| CIN2 | — | 67 (37%) | 35 (39%) | 32 (36%) |
| CIN3 | — | 113 (63%) | 55 (61%) | 58 (64%) |
| **HPV Status at Baseline, n (%)** |  |  |  |  |
| **hrHPV detected** |  |  |  |  |
| Positive | 22 (79%) | 158 (88%) | 81 (90%) | 77 (86%) |
| Negative | 6 (21%) | 22 (12%) | 9 (10%) | 13 (14%) |
| **hrHPV genotype**<br>**(hierarchical)<sup>2</sup>** |  |  |  |  |
| HPV 16 | 6 (21%) | 52 (29%) | 25 (28%) | 27 (30%) |
| HPV 18/45 (without HPV 16) | 4 (14%) | 32 (18%) | 15 (17%) | 17 (19%) |
| HPV 31/33/35/52/58 (without<br>HPV 16, 18/45) | 10 (36%) | 65 (36%) | 35 (39%) | 30 (33%) |
| HPV 51/59 or 39/68/56/66<br>(without HPV 16, 18/45,<br>31/33/35/52/58) | 2 (7%) | 9 (5%) | 6 (7%) | 3 (3%) |
| hrHPV negative | 6 (21%) | 22 (12%) | 9 (10%) | 13 (14%) |
| **Sexually Transmitted Infections at Screening,<sup>3</sup> n (%)** |  |  |  |  |
| **Chlamydia** |  |  |  |  |
| Positive | 0 (0%) | 12 (7%) | 5 (6%) | 7 (8%) |
| **Gonorrhea** |  |  |  |  |
| Positive | 0 (0%) | 6 (3%) | 2 (2%) | 4 (4%) |
| **Trichomonas** |  |  |  |  |
| Positive | 1 (4%) | 19 (11%) | 10 (11%) | 9 (10%) |
| **Syphilis RPR** |  |  |  |  |
| Positive | 1 (3%) | 9 (5%) | 4 (4%) | 5 (6%) |
| **HPV Vaccination, n (%)** |  |  |  |  |
| **Prior HPV vaccination** |  |  |  |  |
| Yes | 1 (3%) | 1 (1%) | 1 (1%) | 0 (0%) |

Q1, 25<sup>th</sup> percentile; Q3, 75<sup>th</sup> percentile; HSIL, high-grade squamous intraepithelial lesion; ASC-H, atypical squamous cells cannot exclude HSIL; LSIL, low-grade squamous intraepithelial lesion; CIN, cervical intraepithelial neoplasia; hrHPV, high-risk human papillomavirus; RPR, rapid plasma reagin.
<sup>1</sup>Participants enrolled under protocol version 4 eligible with either pre-study biopsy or Pap smear.
<sup>2</sup>HPV genotypes are presented hierarchically; participants may be positive for multiple types.
<sup>3</sup>STIs detected at screening were treated prior to study enrollment.
Missing data: HIV viral load missing for 2 randomized participants; pre-study Pap smear missing for 25 randomized participants; pre-study cervical biopsy missing for 40 randomized participants (participants enrolled under amended protocol were eligible with either biopsy or Pap smear).

Most women (96.1%) had high-grade abnormality noted on their pre-study Pap smear, and among those with a pre-study cervical biopsy 41.4% (58/140) had CIN2 and 58.6% (82/140) had CIN3. On baseline LEEP, 37.2% (67/180) had CIN2 and 62.8% (113/180) had CIN3 confirmed. Finally, hrHPV was detected in 87.8% (158/180) of women at baseline. Among the hrHPV-positive women, 32.9% (52/158) had HPV16, 20.3% (32/158) had HPV18/45, and 41.1% (65/158) had HPV31/33/35/52/58 (using hierarchical definitions).

### Acceptability

Acceptability was assessed using a 7-item Likert-scale questionnaire focused on women’s perceptions of ease of use and safety. The questionnaire was administered at Weeks 10 and 24. The proportion of women who found the study cream to be acceptable (score ³ 80%) was high and similar at both timepoints: 95.3% in the 5FU arm vs. 97.8% in the placebo arm at Week 10 (PD -2.4%, 95% CI -9.8%, 3.9%), and 94.0% vs. 94.4% at Week 24 (PD -0.5%, 95% CI -7.4%, 6.5%) (Table 2).

**Table 2.**
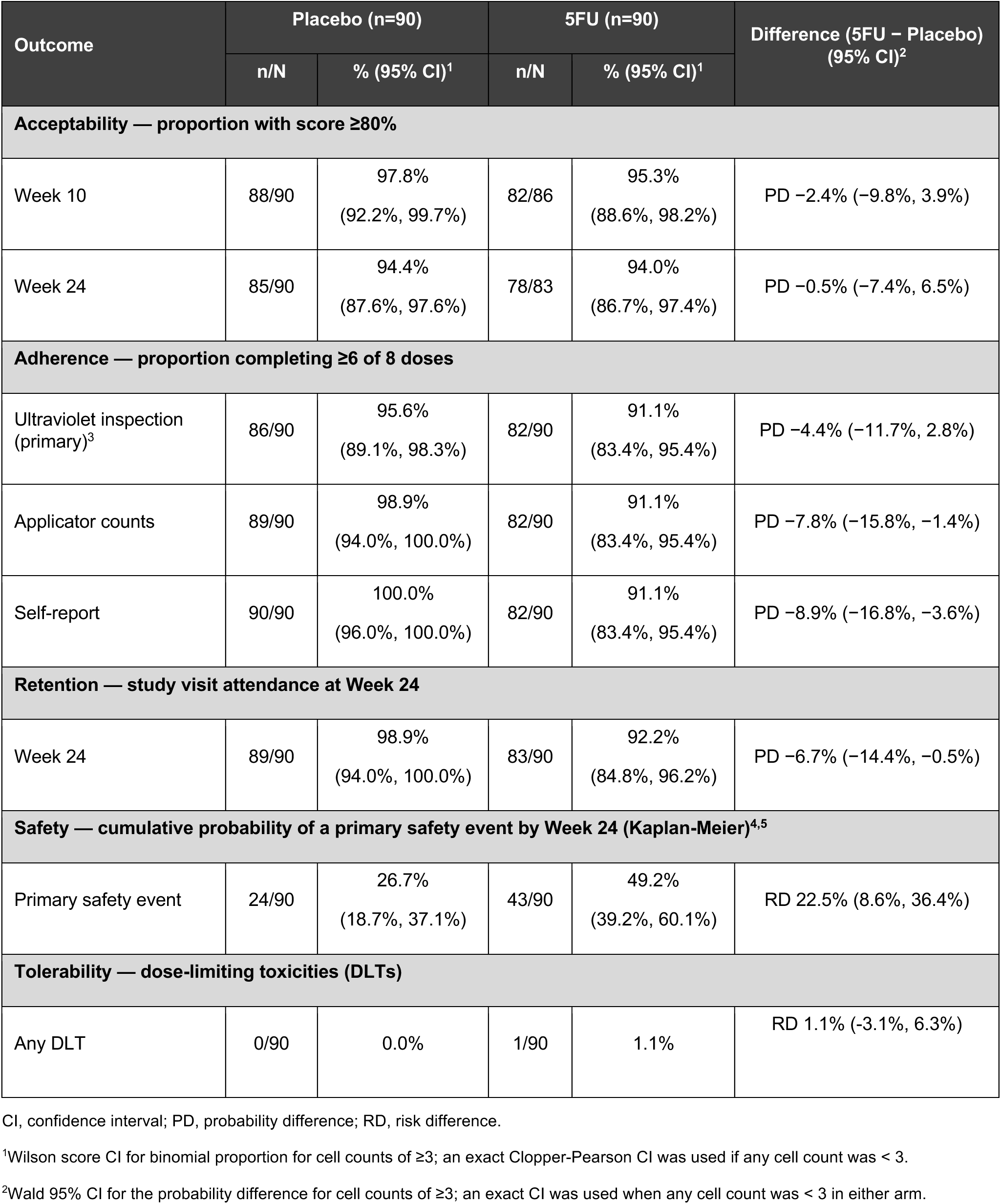
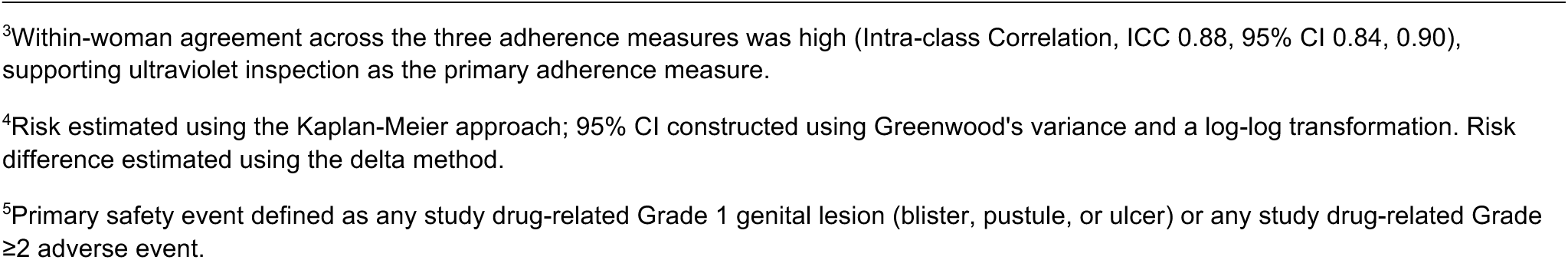
Primary Acceptability and Feasibility Outcomes.

| Outcome | Placebo (n=90) |  | 5FU (n=90) |  | Difference (5FU – Placebo)<br>(95% CI)<sup>2</sup> |
| --- | --- | --- | --- | --- | --- |
|  | n/N | % (95% CI)<sup>1</sup> | n/N | % (95% CI)<sup>1</sup> |  |
| Acceptability — proportion with score ≥80% |  |  |  |  |  |
| Week 10 | 88/90 | 97.8%<br>(92.2%, 99.7%) | 82/86 | 95.3%<br>(88.6%, 98.2%) | PD -2.4% (-9.8%, 3.9%) |
| Week 24 | 85/90 | 94.4%<br>(87.6%, 97.6%) | 78/83 | 94.0%<br>(86.7%, 97.4%) | PD -0.5% (-7.4%, 6.5%) |
| Adherence — proportion completing ≥6 of 8 doses |  |  |  |  |  |
| Ultraviolet inspection (primary)<sup>3</sup> | 86/90 | 95.6%<br>(89.1%, 98.3%) | 82/90 | 91.1%<br>(83.4%, 95.4%) | PD -4.4% (-11.7%, 2.8%) |
| Applicator counts | 89/90 | 98.9%<br>(94.0%, 100.0%) | 82/90 | 91.1%<br>(83.4%, 95.4%) | PD -7.8% (-15.8%, -1.4%) |
| Self-report | 90/90 | 100.0%<br>(96.0%, 100.0%) | 82/90 | 91.1%<br>(83.4%, 95.4%) | PD -8.9% (-16.8%, -3.6%) |
| Retention — study visit attendance at Week 24 |  |  |  |  |  |
| Week 24 | 89/90 | 98.9%<br>(94.0%, 100.0%) | 83/90 | 92.2%<br>(84.8%, 96.2%) | PD -6.7% (-14.4%, -0.5%) |
| Safety — cumulative probability of a primary safety event by Week 24 (Kaplan-Meier)<sup>4,5</sup> |  |  |  |  |  |
| Primary safety event | 24/90 | 26.7%<br>(18.7%, 37.1%) | 43/90 | 49.2%<br>(39.2%, 60.1%) | RD 22.5% (8.6%, 36.4%) |
| Tolerability — dose-limiting toxicities (DLTs) |  |  |  |  |  |
| Any DLT | 0/90 | 0.0% | 1/90 | 1.1% | RD 1.1% (-3.1%, 6.3%) |

CI, confidence interval; PD, probability difference; RD, risk difference.
<sup>1</sup>Wilson score CI for binomial proportion for cell counts of ≥3; an exact Clopper-Pearson CI was used if any cell count was < 3.
<sup>2</sup>Wald 95% CI for the probability difference for cell counts of ≥3; an exact CI was used when any cell count was < 3 in either arm.
<sup>3</sup>Within-woman agreement across the three adherence measures was high (Intra-class Correlation, ICC 0.88, 95% CI 0.84, 0.90), supporting ultraviolet inspection as the primary adherence measure.
<sup>4</sup>Risk estimated using the Kaplan-Meier approach; 95% CI constructed using Greenwood's variance and a log-log transformation. Risk difference estimated using the delta method.
<sup>5</sup>Primary safety event defined as any study drug-related Grade 1 genital lesion (blister, pustule, or ulcer) or any study drug-related Grade $\geq 2$ adverse event.

Median acceptability summary scores were 100% in both arms at both timepoints, with no evident difference between arms at Week 10 (p=0.157) or Week 24 (p=0.852). Across individual items, participants in both arms considered the study cream safe, reported high confidence in their ability to self-administer the cream, and rated their overall experience positively (Supplementary Material).

### Adherence

Adherence was measured using three methods: UVI (primary), applicator counts, and self-report via a dose diary. Within-woman agreement across the three measures was high (ICC 0.88, 95% CI 0.84, 0.90), supporting the use of UVI as the primary measure. By UVI, adherence to ≥6 of 8 doses was 91.1% in the 5FU arm vs. 95.6% in the placebo arm (PD -4.4%, 95% CI -11.7%, 2.8%); this difference was not statistically significant. Adherence estimates were similar by applicator count (Table 2).

### Retention

Retention was high throughout the follow-up period, with 99.4%, 97.8%, 96.1%, and 95.6% of participants attending follow-up visits at Weeks 6, 10, 18, and 24, respectively (Table 2). By Week 24, however, retention differed between study arms, with lower study visit attendance in the 5FU arm compared to the placebo arm (92.2% vs. 98.9%, PD -6.7%, 95% CI -14.4%, -0.5%), as shown in Table 2 and the Supplementary Material. In the 5FU arm, reasons for early study discontinuation included relocation (n=3), loss to follow-up (n=3), and withdrawal of consent (n=1). One participant in the placebo arm relocated and therefore discontinued participation in the study.

### Safety

Study-related safety events were defined as Grade 1 genital lesions (blisters, pustules, or ulcers) or Grade ≥2 AEs that were possibly, probably, or definitely related to the study drug. Overall, 73.3% (132/180) of participants experienced 314 study-related safety events (190 events in the 5FU arm and 124 in the placebo arm). Among these study-related safety events, 4/314 (1.3%) were Grade 1 genital lesions, 95/314 (30.3%) were Grade 2 AEs, and 1/314 (0.3%) were Grade 3 AEs. There were no study-related Grade 4 AEs or deaths.

The probability of experiencing a primary safety event by Week 24 was higher in the 5FU arm than the placebo arm: 49.2% vs. 26.7% (risk difference [RD] 22.5%, 95% CI 8.6%, 36.4%) (Table 2). Using the Kaplan-Meier approach (Supplementary Material), the risk of a primary safety event diverged between arms early in the follow-up and persisted through Week 24. This divergence was driven by Grade 1 and 2 AEs.

### Tolerability

5FU cream was well tolerated in our study population. Only one DLT (a Grade 3 allergic reaction which resolved with treatment) occurred in the 5FU arm, resulting in discontinuation of the study drug. No DLTs occurred in the placebo arm. The participant who experienced the DLT continued study follow-up.

## Discussion

### Main Findings

ACT 2 is the first randomized trial to evaluate a combination treatment approach for CIN2/3 in an LMIC setting. In this randomized feasibility trial, LEEP plus adjuvant intravaginal 5FU was acceptable and feasible among South African WLWH. Acceptability exceeded 94% at both timepoints, and was comparable between arms (PD -0.5%, 95% CI -7.4%, 6.5% at Week 24). Adherence to the study cream (measured by UVI) was also high, exceeding 90% and comparable between arms (PD -4.4%, 95% CI -11.7%, 2.8%). More than 92% of women remained in follow-up throughout the 24-week study period (PD -6.7%, 95% CI -14.4%, -0.5%). These findings support the progression of combination treatment to a Phase 3 efficacy trial.

Our findings of high acceptability and adherence to topical therapies for CIN2/3 are consistent with prior studies in both LMIC^28–31^ and high-income settings.^18, 19^ Building on the lessons from our pre-trial qualitative study^27^ and the vaginal microbicide trial literature,^32–36^ we implemented several strategies to support acceptability and adherence in our study population. These included context-appropriate educational materials; in-clinic coaching and direct observation of the first dose of study cream; illustrated instructions for home use; and text message reminders.

Critically, acceptability remained above 94% and adherence above 91% in the 5FU arm despite a higher burden of AEs compared to placebo. This suggests that women found the every-other-week dosing regimen manageable even when side effects were present, an important finding for future trials, where adherence will be essential to demonstrating treatment benefit. Understanding the nature and frequency of these side effects is also key to designing and implementing the next trial.

Women in the 5FU arm experienced primary safety events nearly twice as frequently as women in the placebo arm (49.2% versus 26.7%). As expected, a large majority of these AEs were Grade 1 or 2 cervical inflammation, consistent with the pharmacological effects of 5FU on the cervical epithelium. Early studies of intravaginal 5FU that used higher-frequency or high-potency dosing regimens resulted in substantially more severe side effects.^37^ Our choice of a lower-potency cream and every-other-week dosing was specifically designed to reduce this burden. Our safety profile is comparable to prior U.S. trials of intravaginal 5FU, in which only Grade 1 and 2 events were observed with every-other-week dosing.^18, 19^

Severe AEs were rare and there were no deaths during the study. One study-related Grade 3 event—an allergic reaction in the 5FU arm—resulted in discontinuation of the study cream. The cause of the allergic reaction was not investigated. However, dihydropyrimidine dehydrogenase (DPD) deficiency, a genetic condition affecting 5FU catabolism, is a known risk factor for severe toxicity to systemic fluoropyrimidines and may be more common in populations of African descent.^38–40^

The difference in retention between the study arms (5FU 92.2% vs. placebo 98.9%) was statistically significant and may be explained by the higher side effect burden in the 5FU arm. This safety signal will therefore need to be accounted for in the design of future Phase 3 trials, including in the informed consent process, monitoring framework, sample size calculations, and attrition assumptions. Pre-screening for DPD deficiency as a safety measure may also be considered, though testing for this deficiency is not readily available in most settings.

### Strengths

Our Phase 2b feasibility trial was rigorously designed to evaluate the practical considerations of studying a new intravaginal intervention for cervical cancer prevention in our setting. Importantly, our double-blind, placebo-controlled design, which is uncommon for feasibility studies, reduced the risk of differential bias in the assessment of outcomes between study arms. The trial sample size was also designed to provide adequate precision and power to assess our primary endpoints, and the precision estimates generated for each outcome provide the foundation for designing a fully powered Phase 3 trial.

Beyond the statistical design, we adapted approaches to study adherence from research on vaginal microbicides, selecting three measures of adherence (applicator counts, UVI, self-report) to minimize the potential for recall and social desirability bias.^41–43^ Finally, safety and tolerability were comprehensively assessed via colposcopy and patient report, using a conservative endpoint definition to capture all events.

### Limitations

Due to the nature of our feasibility design, a major limitation of this trial is the relatively short follow-up of 24 weeks. The trial therefore does not provide data on the durability of our feasibility findings or the longer-term efficacy of the intervention. Additionally, although randomized, our sample size (n=180) was relatively small and our secondary efficacy outcomes (reported elsewhere) were not formally powered.

Our study participants were highly engaged in HIV care and may represent a more adherent group of women than WLWH in the broader region, leading us potentially to overestimate acceptability and adherence compared to the broader population of interest. Moreover, our single-center design (chosen to maximize efficiency) limits the generalizability of our findings to other healthcare settings where the patient population, infrastructure, and implementation context may differ from our specialist, urban hospital.

## Conclusion

ACT 2 is the first randomized controlled trial of LEEP plus adjuvant 5FU in African WLWH and only the second such trial in WLWH worldwide. We demonstrated that this combination treatment approach is acceptable and feasible in the South African setting. Acceptability, adherence, and retention were all high (>90%) and the cream was well tolerated, supporting progression to a larger efficacy evaluation. Future Phase 3 trials should be designed to account for the differential AE burden and retention observed between the study arms.

If proven effective, LEEP plus adjuvant 5FU offers a scalable and pragmatic strategy for improving CIN2/3 treatment outcomes in WLWH. Because 5FU cream is self-administered at home and does not require skilled clinicians, specialized equipment, or clinical infrastructure, this approach is particularly well-suited for low-resource settings, where the burden of cervical disease is greatest.

## Data Availability

Data collection tools and a de-identified dataset are available for research, educational, and non-profit purposes upon request and with appropriate institutional research board or ethics committee approvals.

## Acknowledgements

The authors thank Meghana Mugi, Nhlanhla Kgomari, Tumelo Dlamini, Noluthando Mabandla, Jermina Nkoana, Cassius Dintwe, and Emily Lewis for their invaluable assistance during the trial.

## Contribution to Authorship

CJC and LR conceived and designed the project with input from KRM, NST, JRK, MM, and MF. CJC, NST, MM, SL, TP, MM, and MF participated in participant recruitment and data collection. KRM, JRK, CL, and MM planned and conducted the analyses with input from CJC, NST, and LR. NST and CJC drafted the manuscript and coordinated edits. All authors provided critical input during development of the manuscript and approved the final version. Authors are responsible for the correctness of the statements provided in the manuscript.

## Ethics approval

This study was performed in accordance with the principles of the Declaration of Helsinki. We obtained ethical approval from the University of Witwatersrand Human Research Ethics Committee (201122) and the University of North Carolina at Chapel Hill Institutional Review Board (20-3565). Informed consent was obtained from all participants included in the study.

## Availability of data and materials

Data collection tools and a de-identified dataset are available for research, educational, and non-profit purposes upon request and with appropriate institutional research board and ethics committee approvals.

## Conference presentation

The preliminary results of this study were presented at the 37th Annual Conference of the International Papillomavirus Society (IPVC 2025), Bangkok, Thailand, October 2025.

## REFERENCES

1. Ferlay J EM, Lam F, Laversanne M, Colombet M, Mery L, Piñeros M, Znaor A, Soerjomataram I, Bray F (2024). Global Cancer Observatory: Cancer Today. Lyon, France: International Agency for Research on Cancer. Available from: https://gco.iarc.who.int/today, accessed 10 December 2024. [

2. Bray F, Laversanne M, Sung H, Ferlay J, Siegel RL, Soerjomataram I, et al. Global cancer statistics 2022: GLOBOCAN estimates of incidence and mortality worldwide for 36 cancers in 185 countries. CA Cancer J Clin. 2024;74(3):229–63.

3. Denny L, Anorlu R. Cervical cancer in Africa. Cancer Epidemiol Biomarkers Prev. 2012;21(9):1434–8.

4. Stelzle D, Tanaka LF, Lee KK, Ibrahim Khalil A, Baussano I, Shah ASV, et al. Estimates of the global burden of cervical cancer associated with HIV. Lancet Glob Health. 2021;9(2):e161–e9.

5. Saslow D, Solomon D, Lawson HW, Killackey M, Kulasingam SL, Cain J, et al. American Cancer Society, American Society for Colposcopy and Cervical Pathology, and American Society for Clinical Pathology Screening Guidelines for Prevention and Early Detection of Cervical Cancer. J Low Genit Tract Dis. 2012;16:175–204.

6. Katki HA, Schiffman M, Castle PE, Fetterman B, Poitras NE, Lorey T, et al. Five-year risk of recurrence after treatment of CIN 2, CIN 3, or AIS: performance of HPV and Pap cotesting in posttreatment management. J Low Genit Tract Dis. 2013;17:S78–84.

7. American College of Obstetricians and Gyncologists. Management of abnormal cervical cancer screening test results and cervical cancer precursors. Practice Bulletin No. 140. Obstet Gynecol. 2013;122:1338–67.

8. Heard I. Prevention of cervical cancer in women with HIV. Current opinion in HIV and AIDS. 2009;4(1):68–73.

9. Reimers LL, Sotardi S, Daniel D, Chiu LG, Van Arsdale A, Wieland DL, et al. Outcomes after an excisional procedure for cervical intraepithelial neoplasia in HIV-infected women. Gynecologic oncology. 2010;119(1):92–7.

10. Massad LS, Fazzari MJ, Anastos K, Klein RS, Minkoff H, Jamieson DJ, et al. Outcomes after treatment of cervical intraepithelial neoplasia among women with HIV. Journal of lower genital tract disease. 2007;11(2):90–7.

11. Smith JS, Sanusi B, Swarts A, Faesen M, Levin S, Goeieman B, et al. A randomized clinical trial comparing cervical dysplasia treatment with cryotherapy vs loop electrosurgical excision procedure in HIV-seropositive women from Johannesburg, South Africa. Am J Obstet Gynecol. 2017.

12. Greene SA, De Vuyst H, John-Stewart GC, Richardson BA, McGrath CJ, Marson KG, et al. Effect of Cryotherapy vs Loop Electrosurgical Excision Procedure on Cervical Disease Recurrence Among Women With HIV and High-Grade Cervical Lesions in Kenya: A Randomized Clinical Trial. JAMA. 2019;322(16):1570–9.

13. Firnhaber C, Swarts A, Jezile V, Mulongo M, Goeieman B, Williams S, et al. Human Papillomavirus Vaccination Prior to Loop Electroexcision Procedure Does Not Prevent Recurrent Cervical High-grade Squamous Intraepithelial Lesions in Women Living With Human Immunodeficiency Virus: A Randomized, Double-blind, Placebo-controlled Trial. Clin Infect Dis. 2021;73(7):e2211–e6.

14. Debeaudrap P, Sobngwi J, Tebeu PM, Clifford GM. Residual or Recurrent Precancerous Lesions After Treatment of Cervical Lesions in Human Immunodeficiency Virus-infected Women: A Systematic Review and Meta-analysis of Treatment Failure. Clin Infect Dis. 2019;69(9):1555–65.

15. Stanley M. Chapter 17: Genital human papillomavirus infections--current and prospective therapies. J Natl Cancer Inst Monogr. 2003(31):117–24.

16. van de Nieuwenhof HP, van der Avoort IA, de Hullu JA. Review of squamous premalignant vulvar lesions. Crit Rev Oncol Hematol. 2008;68(2):131–56.

17. Weis SE. Current treatment options for management of anal intraepithelial neoplasia. Onco Targets Ther. 2013;6:651–65.

18. Maiman M, Watts DH, Andersen J, Clax P, Merino M, Kendall MA. Vaginal 5-fluorouracil for high-grade cervical dysplasia in human immunodeficiency virus infection: a randomized trial. Obstet Gynecol. 1999;94(6):954–61.

19. Rahangdale L, Lippmann QK, Garcia K, Budwit D, Smith JS, van Le L. Topical 5-fluorouracil for treatment of cervical intraepithelial neoplasia 2: a randomized controlled trial. Am J Obstet Gynecol. 2014;210(4):314 e1-8.

20. Chibwesha CJ, Mollan KR, Teodoro NS, Keys JR, Liu C, Mulongo M, et al. Randomized clinical trial protocol: Acceptability and feasibility of combination treatment for cervical precancer among South African women living with HIV (ACT 2). Contemp Clin Trials. 2026;164:108272.

21. Mbulawa ZZA, Wilkin TJ, Goeieman B, Swarts A, Williams S, Levin S, et al. Xpert human papillomavirus test is a promising cervical cancer screening test for HIV-seropositive women. Papillomavirus Res. 2016;2:56–60.

22. Jacobsson S, Boiko I, Golparian D, Blondeel K, Kiarie J, Toskin I, et al. WHO laboratory validation of Xpert((R)) CT/NG and Xpert((R)) TV on the GeneXpert system verifies high performances. APMIS. 2018;126(12):907–12.

23. Gaydos CA, Van Der Pol B, Jett-Goheen M, Barnes M, Quinn N, Clark C, et al. Performance of the Cepheid CT/NG Xpert Rapid PCR Test for Detection of Chlamydia trachomatis and Neisseria gonorrhoeae. J Clin Microbiol. 2013;51(6):1666–72.

24. Garrett N, Mitchev N, Osman F, Naidoo J, Dorward J, Singh R, et al. Diagnostic accuracy of the Xpert CT/NG and OSOM Trichomonas Rapid assays for point-of-care STI testing among young women in South Africa: a cross-sectional study. BMJ open. 2019;9(2):e026888.

25. Schwebke JR, Gaydos CA, Davis T, Marrazzo J, Furgerson D, Taylor SN, et al. Clinical Evaluation of the Cepheid Xpert TV Assay for Detection of Trichomonas vaginalis with Prospectively Collected Specimens from Men and Women. J Clin Microbiol. 2018;56(2).

26. Nakku-Joloba E, Kiragga A, Mbazira JK, Kambugu F, Jett-Goheen M, Ratanshi RP, et al. Clinical Evaluation of 2 Point-of-Care Lateral Flow Tests for the Diagnosis of Syphilis. Sex Transm Dis. 2016;43(10):623–5.

27. Teodoro NS, Ngcobo N, Jean-Baptiste M, Mulongo M, Milford C, Beksinska M, et al. Perspectives on Cervical Cancer Prevention Strategies and a Combination Treatment for Cervical Precancer in South African Women Living with HIV and Male Partners. medRxiv. 2025.

28. Mungo C, Kachoria AG, Adoyo E, Zulu G, Goraya SK, Omoto J, et al. ’ARVs is for HIV and cream is for HPV or precancer:’ Women’s perceptions and perceived acceptability of self-administered topical therapies for cervical precancer treatment: a qualitative study from Kenya. Ecancermedicalscience. 2025;19:1903.

29. Adewumi K, Kachoria AG, Adoyo E, Rop M, Owaya A, Tang JH, et al. Women’s experiences and acceptability of self-administered, home delivered, intravaginal 5-Fluorouracil cream for cervical precancer treatment in Kenya. Front Reprod Health. 2025;7:1487264.

30. Mungo C, Adewumi K, Ellis G, Rop M, Adoyo E, Zou Y, et al. Men’s perceptions and perceived acceptability of their female partner’s use of self-administered intravaginal therapies for treatment of cervical precancer in Kenya. Ecancermedicalscience. 2024;18:1719.

31. Mungo C, Adewumi K, Adoyo E, Zulu G, Goraya SK, Ogollah C, et al. “There is nothing that can prevent me from supporting her:” men’s perspectives on their involvement and support of women’s use of topical therapy for cervical precancer treatment in Kenya. Front Oncol. 2024;14:1360337.

32. van der Straten A, Van Damme L, Haberer JE, Bangsberg DR. Unraveling the divergent results of pre-exposure prophylaxis trials for HIV prevention. AIDS. 2012;26(7):F13–9.

33. Minnis AM, Gandham S, Richardson BA, Guddera V, Chen BA, Salata R, et al. Adherence and acceptability in MTN 001: a randomized cross-over trial of daily oral and topical tenofovir for HIV prevention in women. AIDS Behav. 2013;17(2):737–47.

34. MacQueen KM, Weaver MA, van Loggerenberg F, Succop S, Majola N, Taylor D, et al. Assessing adherence in the CAPRISA 004 tenofovir gel HIV prevention trial: results of a nested case-control study. AIDS Behav. 2014;18(5):826–32.

35. Kashuba AD, Gengiah TN, Werner L, Yang KH, White NR, Karim QA, et al. Genital Tenofovir Concentrations Correlate With Protection Against HIV Infection in the CAPRISA 004 Trial: Importance of Adherence for Microbicide Effectiveness. J Acquir Immune Defic Syndr. 2015;69(3):264–9.

36. Abdool Karim Q, Abdool Karim SS, Frohlich JA, Grobler AC, Baxter C, Mansoor LE, et al. Effectiveness and safety of tenofovir gel, an antiretroviral microbicide, for the prevention of HIV infection in women. Science. 2010;329(5996):1168–74.

37. Krebs HB, Helmkamp BF. Chronic ulcerations following topical therapy with 5-fluorouracil for vaginal human papillomavirus-associated lesions. Obstet Gynecol. 1991;78(2):205–8.

38. Amstutz U, Froehlich TK, Largiader CR. Dihydropyrimidine dehydrogenase gene as a major predictor of severe 5-fluorouracil toxicity. Pharmacogenomics. 2011;12(9):1321–36.

39. Kishi P, Price CJ. Life-Threatening Reaction with Topical 5-Fluorouracil. Drug Saf Case Rep. 2018;5(1):4.

40. Mattison LK, Fourie J, Desmond RA, Modak A, Saif MW, Diasio RB. Increased prevalence of dihydropyrimidine dehydrogenase deficiency in African-Americans compared with Caucasians. Clin Cancer Res. 2006;12(18):5491–5.

41. Curran K, Mugo NR, Kurth A, Ngure K, Heffron R, Donnell D, et al. Daily short message service surveys to measure sexual behavior and pre-exposure prophylaxis use among Kenyan men and women. AIDS Behav. 2013;17(9):2977–85.

42. Pietanza MC, Basch EM, Lash A, Schwartz LH, Ginsberg MS, Zhao B, et al. Harnessing technology to improve clinical trials: study of real-time informatics to collect data, toxicities, image response assessments, and patient-reported outcomes in a phase II clinical trial. J Clin Oncol. 2013;31(16):2004–9.

43. Andriesen J, Bull S, Dietrich J, Haberer JE, Van Der Pol B, Voronin Y, et al. Using Digital Technologies in Clinical HIV Research: Real-World Applications and Considerations for Future Work. J Med Internet Res. 2017;19(7):e274.

